# Theory of Mind Imitation by LLMs for Physician-Like Human Evaluation

**DOI:** 10.1101/2025.03.01.25323142

**Authors:** Raghav Awasthi, Shreya Mishra, Charumathi Raghu, Moises Auron, Ashish Atreja, Dwarikanath Mahapatra, Nishant Singh, Ashish K. Khanna, Jacek B. Cywinski, Kamal Maheshwari, Francis A. Papay, Piyush Mathur

## Abstract

Aligning the Theory of Mind (ToM) capabilities of Large Language Models (LLMs) with human cognitive processes enables them to imitate physician behavior. This study evaluates LLMs abilities such as Belief and Knowledge, Reasoning and Problem-Solving, Communication and Language Skills, Emotional and Social Intelligence, Self-Awareness, and Metacognition in performing human-like evaluations of Foundation Models. We used a dataset composed of clinical questions, reference answers, and LLM-generated responses based on guidelines for the prevention of heart disease. Comparing GPT-4 to human experts across ToM abilities, we found the highest Emotional and Social Intelligence agreement using the Brennan-Prediger coefficient. This study contributes to a deeper understanding of LLM’s cognitive capabilities and highlights their potential role in augmenting or complementing human clinical assessments.

## Introduction

Theory of Mind refers to the ability of the human mind to attribute mental states to others, which large language models have tried to emulate. Many of the recent experiments in healthcare have focused on testing the ability of LLMs to assess if they can reason like a physician. Technical and human evaluations have been performed to assess the LLM’s capabilities of inferring other states of mind and matching human values. Both technical and human evaluations have their strength and limitations to evaluate the veracity of LLM performance in a specialized field such as medicine. However, human evaluation is considered essential to assuring safety and effectiveness. Multiple human evaluation frameworks have recently been proposed, covering key aspects of model evaluation such as relevance, coverage, harm, and coherence.(Elangovan et al. 2024; Tam et al. 2024; Awasthi et al. 2023). These key aspects involve deep human cognitive processes, encompassing ToM abilities such as Belief and Knowledge, Reasoning and Problem-Solving, Communication and Language Skills, Emotional and Social Intelligence, Self-Awareness, and Metacognition, making human evaluation more trustworthy. However, human evaluations at scale are expensive and time-consuming requiring coordination with annotators, custom interfaces, and detailed instructions (Elangovan et al. 2024; Hosking, Blunsom, and Bartolo 2023). To overcome these challenges, recent solutions have proposed using LLMs as an alternative to human evaluation (Chiang and Lee 2023). However, studies have not evaluated LLMs for human evaluation under Theory of Mind abilities, especially with the key social metrics such as biasness, toxicity and privacy(Bedi et al. 2024) We hypothesize that LLMs have the ability to infer similar to human evaluation using metrics which are highly correlated with ToM abilities (Chen et al. 2024). We experiment with GPTs performance across diverse metrics representing different contexts of evaluations, including important social ones to test the ability of LLMs to evaluate like a physician.

## Methods

### Data

Clinical experts developed a question-answer dataset based on the 2019 AHA Guideline on the Primary Prevention of Cardiovascular Disease (Arnett et al. 2019). Model answers were generated using GPT-4o (Achiam et al. 2023), and LLaMA-3 ((7B Chat)) (Touvron et al. 2023) in Retrieval Augmented Generation (RAG) and No RAG setup. In the RAG setup, the original guideline was provided as context. Three reviewers evaluated the generated answers using a robust human evaluation framework with 15 metrics covering Relevance (accuracy, comprehension, reasoning, helpfulness), Coverage (key points, retrieval, missingness), Coherence (fluency, grammar, organization), and Harm (bias, toxicity, privacy, hallucination). Ratings were based on a 1–5 Likert scale, where 1 indicated strong disagreement and five strong agreement. Next, we used GPT-4 with a zero-shot prompt to generate the ratings for all 15 metrics.

### Mapping ToM Abilities with Human Evaluation Metrics

We have mapped HumanELY (Awasthi et al. 2023) based 15 human evaluation metrics into five different **(Figure 1 A)** categories of ToM capabilities, including Belief and Knowledge, Reasoning and Problem-Solving, Communication and Social Intelligence, and Self-Awareness and Metacognition (Chen et al. 2024).

**Figure 1:**
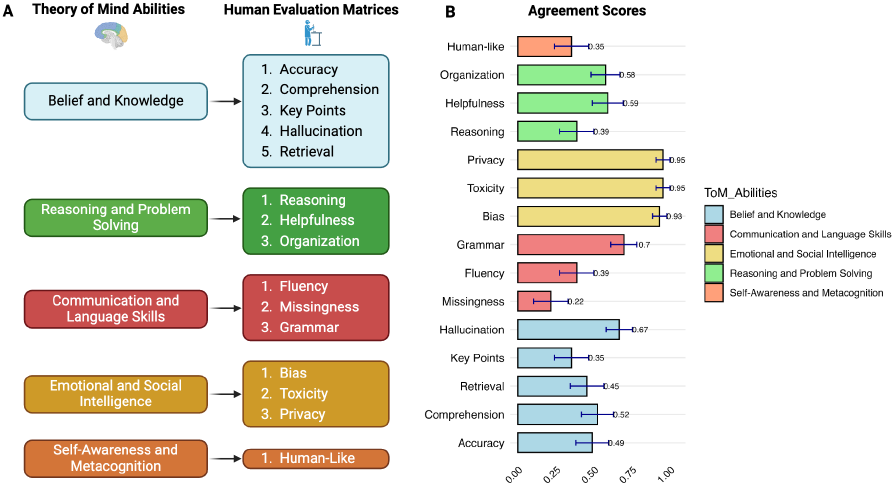
A) Mapping of 15 human evaluation metrics into five categories of ToM abilities. B) Bar plot of Brennan-Prediger’s (BP) agreement coefficient scores, with standard error represented as error bars, color-coded based on ToM categories.

### Analysis

We compared GPT-4 ratings with human reviewers using the Brennan-Prediger (BP) agreement coefficient (Moss 2023) across all ToM abilities. First, we obtained the consensus of the three human reviewers by taking the majority vote. Then, we grouped ratings into broader categories, combining 1 and 2 (disagreement) and 4 and 5 (agreement) to assess the LLM’s alignment with human consensus, focusing on agreement, disagreement, and neutrality. Finally, human consensus ratings were compared to GPT-4 ratings. Agreement scores are reported with standard error and p-values.

## Results

### ToM Ability 1: Beliefs and Knowledge

From the Brennan-Prediger’s (BP) agreement coefficient **(Figure 1 B)**, we found that metrics associated with beliefs and knowledge demonstrated varying levels of agreement. Accuracy achieved a moderate agreement score of 0.49 (SE = 0.11, p-value *<*0.05), while Comprehension scored slightly higher at 0.52 (SE = 0.11, p-value *<*0.05). Retrieval achieved a BP score of 0.45 (SE = 0.11, p-value *<*0.05), indicating moderate alignment in identifying and retrieving relevant information. However, Key Points scored 0.35 (SE = 0.11, p-value *<*0.05), suggesting less consensus in capturing critical elements. Hallucination demonstrated substantial alignment, achieving a BP score of 0.67 (SE = 0.09, p-value *<*0.05), reflecting the model’s ability to maintain factual consistency.

### ToM Ability 2: Reasoning and Problem-Solving

Metrics related to reasoning and problem-solving achieved moderate agreement levels. Reasoning had a BP score of 0.39 (SE = 0.11, p-value *<*0.05), reflecting limited alignment in evaluations. Helpfulness scored higher, with a BP score of 0.59 (SE = 0.10, p-value *<*0.05), indicating more robust agreement. The organization demonstrated moderate alignment, achieving a BP score 0.58 (SE = 0.09, p-value *<*0.05). These results highlight that while LLMs exhibit some capacity for problem-solving and reasoning.

### ToM Ability 3: Communication and Language Skills

Communication and language-related metrics varied widely in agreement levels. Grammar achieved the highest score in this category, with a BP score of 0.70 (SE = 0.09, p-value *<*0.05), indicating substantial alignment. Fluency scored 0.39 (SE = 0.11, p-value *<*0.05), reflecting modest alignment. Conversely, Missingness had the lowest score in this category (BP = 0.22, SE = 0.11, p-value *<*0.05), suggesting limited consensus on identifying missing information.

### ToM Ability 4: Emotional and Social Intelligence

Metrics assessing emotional and social intelligence demonstrated the highest levels of agreement across all categories. Bias achieved a near-perfect BP score of 0.93 (SE = 0.05, p-value *<*0.05), alongside Privacy and Toxicity, scoring 0.95 (SE = 0.05, p-value *<*0.05). These results indicate that LLMs could be helpful in annotating harm-related factors.

### ToM Ability 5: Self-Awareness and Metacognition

Self-awareness and metacognition metrics showed modest agreement, with human-like responses scoring 0.35 (SE = 0.11, p-value *<*0.05). This indicates moderate agreement with human experts in this cognitive domain.

## Discussion & Conclusion

Researchers have struggled with evaluation of ToM based AI algorithms as human preferences for these are sometimes vague and hard to specify (Langley et al. 2022). Results of our experiments highlight strong agreement for metrics of harm, with moderate to lower alignment for relevance, coverage, and coherence metrics, pointing to areas for further refinement in evaluation consistency. LLMs possibly have the potential to overcome the barrier of unscalable human evaluation, especially for ToM social metrics aligned with harm in healthcare, and require inferring human thinking for assessment. To evaluate LLMs performance in healthcare and ability to reason like a physician, all the three approaches for implementation of AI, data-centric, model-centric and human-centric, must converge. LLMs themselves can possibly have significant capabilities to emulate physician-like state of mind to perform evaluations providing us with the ability to perform scalable, measurable, comparable and comprehensive evaluations. Such evaluations would then match assessment of all the three key approaches of AI implementation in healthcare, build trust and escalate adoption of LLMs in healthcare. We also hope that our experimentations provide the much needed signal to further refine ToM based AI algorithms and automate robust evaluations(Shah et al. 2021; Langley et al. 2022).

### Limitation

We acknowledge that these experiments are based on a single dataset, which limits the generalizability of our findings.

## Data Availability

All data produced in the present study are available upon reasonable request to the authors.

